# Social distancing and preventive practices of government employees in response to COVID-19 in Ethiopia

**DOI:** 10.1101/2020.12.15.20248271

**Authors:** Wakgari Deressa, Alemayehu Worku, Workeabeba Abebe, Sefonias Getachew, Wondwosson Amogne

**Affiliations:** Department of Preventive Medicine, School of Public Health, College of Health Sciences, Addis Ababa University, Addis Ababa, Ethiopia; Department of Pediatrics and Child Health, School of Medicine, College of Health Sciences, Addis Ababa University, Addis Ababa, Ethiopia; Department of Internal Medicine, School of Medicine, College of Health Sciences, Addis Ababa University, Addis Ababa, Ethiopia

**Keywords:** COVID-19, Ethiopia, Facemask, Government employees, SARS-COV-2, Social distancing

## Abstract

**Background:** Public health measures are critical to mitigate the spread of the novel coronavirus disease 2019 (COVID-19) pandemic. Ethiopia has implemented a variety of essential public health measures in response to the spread of the virus. This study aimed to assess social distancing and preventive practices of government employees in response to COVID-19.

**Methods:** A cross-sectional study was conducted among 1573 government employees selected from 46 public institutions (16 National, 18 from Addis Ababa City Administration, and 12 from Oromia Regional State) located in Addis Ababa. Data were collected from 8^th^ to 19^th^ June 2020 using a paper-based self-administered questionnaire and analyzed using SPSS version 23.0. ANOVA and t-tests were applied to assess the difference between groups. Bivariate and multivariable logistic regression analyses were used to identify factors associated with outcome variables.

**Results:** The majority of the participants reported wearing of facemask (96%), avoiding close contact with people including handshaking (94.5%), frequent had washing (94.1%), maintaining physical distancing (89.5%), avoiding mass gatherings (88.1%), and restricting movement and travelling (84.1%). More than 80% of the participants perceived that consistently wearing a facemask is highly effective in preventing the transmission of coronavirus. Participants from Oromia reported statistically significantly lower odds of perceived effectiveness of facemask in preventing coronavirus infection (adjusted OR=0.27, 95% CI:0.17-0.45). About 19% of the respondents reported that they had ever tested for COVID-19. Participants within the age groups of 18-29 were more likely to test for coronavirus than the older age groups. Whilst, respondents from Oromia were less likely to test for coronavirus than those from national level (adjusted OR=0.31, 95% CI:0.16-0.60). About one-third (31.3%) of the respondents strongly agreed that the policy responses that the Government had taken to contain the spread of coronavirus were reasonable, and 38.5% agreed with the policy responses.

**Conclusions:** The findings showed higher social distancing and preventive practices among the government employees in response to COVID-19. People should properly apply social distancing measures, wearing facemasks, and washing hands frequently with water and soap as a comprehensive package of SARS-CoV-2 prevention and control strategies. Rules and regulations imposed by the Government should be properly enforced in order to control the pandemic.

## Background

The ongoing rapid spread of the novel coronavirus disease 2019 (COVID-19), caused by severe acute respiratory syndrome coronavirus 2 (SARS-CoV-2) [1] and first reported in China in December 2019 [2, 3], has led the World Health Organization (WHO) to declare the disease as a global pandemic of public health threat on 11^th^ March 2020 [4]. Since then, the virus has continually been spread across the world at an extreme rate causing unprecedented deaths mainly among older age people with underlying health conditions like chronic lung disease, hypertension, cardiovascular disease, heart disease, diabetes, and obesity [5, 6]. This extraordinary rapid transmission of the virus has already reached at least 220 countries and territories around the world and caused over 72 million confirmed cases and more than 1.6 million deaths worldwide as of 14^th^ December 2020 [7]. Almost all African countries have been hit with the pandemic with the first confirmed case reported in Egypt on 14^th^ February 2020 [8]. As of 14^th^ December 2020, more than 2.3 million confirmed COVID-19 cases with 56,440 deaths have been reported from Africa, with the majority of cases reported from South Africa, Morocco, Egypt, Ethiopia and Tunisia [7].

Ethiopia reported its first confirmed case of COVID-19 on 13^th^ March 2020 [9]. The index case was a foreigner who tested positive by the Ethiopian Public Health Institute (EPHI). Three new secondary cases that were linked to the index case and an additional confirmed COVID-19 case imported from Dubai were reported on 15^th^ March. Since the report of the index case, updates and press statements on the situation of the pandemic in the country have been daily given to the public by the Ministry of Health (MoH) and EPHI. Most of the cases during the early phase of the pandemic were detected among people with travel history of abroad, mandatory quarantined passengers, and health screening at the points of entry to the country [10]. Very soon, new cases were reported among close contacts of the confirmed cases, and the disease then quickly spread to the community. The health authorities have since early February stepped up various prevention and intervention activities against COVID-19. Extensive contact tracing and immediate isolation of the confirmed cases was implemented by the Government during the early stages of the pandemic. However, within less than three months after the index case of COVID-19, the virus quickly spread to all parts of the country. By the first week of June, all regions reported COVID-19 cases, with Addis Ababa constituting about 75% of the cases [11]. Increased number of imported cases along with increased number of secondary cases subsequently contributed to community transmission. As of 13^th^ December, 2020, the number of confirmed COVID-19 cases in Ethiopia has reached 116,769 and the number of deaths is 1,806.

In the absence of specific therapeutics or effective immunization particularly during the early stages of a potentially pandemic outbreak such as COVID-19, public health measures are critical to prevent and interrupt the person-to-person transmission of the virus through respiratory droplets and close contact [12]. In order to reduce or contain the spread of SARS-COV-2 and its associated mortality rates, many countries have implemented a lot of public health measures such as isolation, quarantine, social distancing, facemask wearing and hand hygiene practices [13-15], and these measures have proved to be effective in many countries [16-19]. Mathematical models of COVID19 spread have demonstrated the impact of social distancing [20] and universal masking [21] on the reduction of the spread of coronavirus. Earlier studies on the global outbreak of severe acute respiratory syndrome (SARS) also demonstrated that the spread of pandemic influenza was substantially reduced by diligent hand hygiene practices and mask wearing [22, 23]. This implies that frequent handwashing with soap and water, wearing facemask, social distancing and avoiding close contacts with other people are the simplest measures that can be applied by everyone to protect themselves from COVID-19.

The initial containment measures used to contain COVID-19 in Ethiopia during March and April 2020 included intense surveillance for infections, not only in incoming travelers but also screening of individuals at high risk of infection who had close contact with a confirmed case, immediate isolation of all confirmed cases, quarantine, and a public campaign for social distancing and preventive practices. While many governments around the world have implemented drastic measures to slow down the rate of transmission of COVID-19 such as severe travel restrictions and lockdowns [24], the Government of Ethiopia has implemented a variety of less drastic essential measures in response to the spread of the virus, such as airport surveillance and suspension of flights, travel restrictions, closure of international borders, flexible working arrangements, closing schools and universities, and mandatory quarantine well ahead of many countries around the world. Religious organizations cancelled services from March 31^st^ onwards, and conferences and other mass gatherings and sports have been banned. State of emergency was declared on 8^th^ April 2020 [25]. At the end of May, wearing facemask in public was enforced as mandatory.

The current public health interventions highly promoted and implemented in Ethiopia as part of the efforts to control the rate of transmission of the SARS-CoV-2 at individual level include frequent handwashing with soap and water, social distancing, wearing facemask including home-made masks, use of alcohol-based sanitizers, staying at home when possible, covering mouth and nose when coughing and sneezing, and not touching the nose, mouth and eyes, and refraining from risky behaviors such as travel and mass gatherings. Continuous investigation through laboratory testing, case detection, isolation and contact tracing has been the milestone of the control efforts throughout the country to better understand the transmission dynamics and strengthen appropriate prevention and control strategies [26]. Diagnostic testing capacity was scaled-up from zero in early March to over 8,000 tests per day by early August 2020. Despite all the public health measures, the daily number of positive cases is steadily increasing in the country, but at a slower rate than earlier estimates.

Studies have shown that strong public health measures such as social distancing and preventive behaviors have resulted in a substantial reduction in the transmission of COVID-19 [16, 20]. The impact of public health interventions and population behavioral changes that have been rolled out in Ethiopia to contain COVID-19 transmission has not been evaluated. High public compliance to proper risk reduction measures such as practicing social distancing, wearing mask, frequent handwashing and staying home can be effectively achieved if the public understands and is persuaded of the importance of these measures in the prevention and control of COVID-19 [27]. Thus far, very limited research has reported on how individuals have practiced protective behaviors in response to COVID-19 pandemic in Ethiopia [28-30]. The aim of this study was to assess the health protective measures such as social distancing and preventive behaviors of government employees in Addis Ababa in response to COVID-19. The results of this study are important to inform future efforts focusing on the people’s readiness to comply with pandemic control measures and the development of preventive strategies and health promotion programs, given that proper practices of social distancing and preventive behaviors can play important roles in the prevention and control of COVID-19.

## Methods

### Study setting

This study was conducted in Addis Ababa city administration three months after the first confirmed COVID-19 case in Ethiopia. Addis Ababa city has the highest rate of COVID-19 cases and deaths in Ethiopia, and is considered as an epicenter of COVID-19 in the country. Of the total 4,070 confirmed COVID-19 cases reported in the country as of 19^th^ June, the majority (73.4%) of the cases were reported from Addis Ababa. During the data collection period between 8^th^ and 19^th^ June 2020, the total number of confirmed new COVID-19 cases reported in the country was 2,064 including 45 deaths, of which Addis Ababa contributed 71.6% of the cases and 89% of the deaths. During the 12 days of data collection, the number of COVID-19 in Addis Ababa increased from 1,625 on 8^th^ June to 2,988 on 19^th^ June, representing an increase of 84%. The most impacted sub-cities included Addis Ketema, Lideta, and Gulele, while Akaki sub-city had the lowest number of cases, and most of the cases were due to community transmission as of the first week of May.

### Study design and sampling

A cross-sectional survey was conducted among government employees of 46 public institutions located in Addis Ababa. Due to physical distancing restrictions, it was not possible to conduct a representative community-based face-to-face interview during this period. As a result, this study collected data using institution-based self-administered survey. The study population for this study constituted all government employees, working in the selected government institution at the time of the survey and willing to participate in the study. These included professionals, experts, technicians and support staff working at different hierarchies and divisions/directorates in the institution including higher and midlevel officials.

A sample size of 1,710 was calculated with a precision of 4% to estimate a 50% proportion with 95% confidence, a design effect of 2 and 30% non-response rate. Purposive sampling was used to select public institutions located in Addis Ababa city administration. The institutions were stratified into three government levels and selected from the national or Federal Government Ministries, Addis Ababa city administration bureaus and sub-cities, and Oromia Regional State bureaus located in Addis Ababa [Additional file 1]. The study participants were assumed to represent employees from the community in Addis Ababa and its environs involved in policy and decision-making processes. The decisions and practices made by these people would subsequently have direct or indirect influence on individuals, family and the community in response to COVID-19.

The data collectors initially contacted the respective higher official in the institution to explain the purpose of the survey and submit the support letter. After approval of the support letter, the Human Resource Directorate of the respective institutions were contacted to obtain information on the total number of employees, number of directorates and departments in the institution with their respective number of personnel. The sample size initially allocated to the institution was distributed to the directorates or departments proportional to the size of their employees. Emphasis was given to equally select the participants, to proportionally distribute the number of questionnaires to the different directorates or departments in the selected institutions based on the size of their employees. In this survey, we tried to avoid selection bias by including as many representative respondents’ as possible within the shortest possible time. Since some employees were working on a shift basis due to the current situation of COVID-19 pandemic, their availability was taken into consideration while distributing the questionnaires. When the selected respondent was known that he/she couldn’t return to the office during the first 2-3 days of the survey, replacement was made. Emphasis was also given to ensure the gender balance during the selection of the respondents and distribution of the questionnaires.

### Data collection

A paper-based self-administered questionnaire was used to collect data [Additional file 2]. The questionnaire was developed by the research team for the purpose of this survey, and some questions were adapted from the WHO tools used for a similar study [31]. The questionnaire had three main parts: (1) socio-demographic characteristics; (2) social distancing, and (3) preventive practices. The tool was initially developed in English and translated into *Amharic* and *Afan Oromo* by experienced personnel, and back translated into English for accuracy by independent personnel. Trained personnel with previous experience collected the data using standardized paper-based self-administered questionnaires. The questionnaire included an introductory information on the cover letter to inform participants about the study and explaining the purpose of the survey, consent information to ensure voluntary participation in the study while ensuring confidentiality of data, and researchers contact information for any questions the respondent might have. Individuals who declined to participate were excluded from the survey. Participants completed the questionnaires by themselves in the local language (*Afan* Oromo in the Oromia Regional State Offices and *Amharic* otherwise).

In this study, social distancing and preventive practices were defined as the main health protective measures that are adopted and applied by people to protect themselves and others from contracting disease pandemics such as COVID-19 and slowing down the spread of the virus [32-34]. Social distancing practices include physical distancing, staying at home when sick, working from work, avoiding mass gatherings, social events, crowded places, public transport and travelling, and avoiding close contact with people including shaking hands or hugging. Physical distancing involves the practice of maintaining at least two adult strides or two meters distance between two or more people. Preventive behaviors or hygiene practices include wearing a facemask, washing hands more frequently with water and soap, using hand sanitizer more regularly, cleaning and disinfecting surfaces including mobile phones, avoiding touching eyes, nose and mouth, and covering the mouth and nose when coughing and sneezing using a tissue paper or bent elbow.

### Statistical analyses

Data were entered into the Census and Survey Processing System (CSPro) software package, version 7.2 (U.S. Census Bureau and ICF Macro) and analyzed using Statistical Package for Social Sciences (SPSS) version 23 (SPSS Inc., IBM, USA). The outcome of interest was COVID-19 related protective practices (social distancing and preventive practices) taken by individuals. These variables were based on the question “Which of the following measures, if any, are you currently taking to prevent yourself against COVID-19”? Respondents were able to select from 14 possible protective health measures including staying at home, maintaining physical distancing, avoiding close contact with people including handshaking, covering mouth/nose with face/cloth mask when going outdoors, washing hands with soap and water frequently, avoiding touching eyes, nose and mouth, avoiding mass gathering, covering mouth/nose while coughing or sneezing, restricting movement, testing for novel coronavirus, recommending the use of facemask to people when going outdoors, and following government recommendations to combat COVID-19. Responses were recoded as ‘1=Yes’ and ‘0=No’. A composite index of the average of all items was created for each respondent to form total preventive measures being taken by the individual, ranging from 0 to 14, with a higher score indicating that participants demonstrate higher protective measures. The internal consistency of the items was moderate (Cronbach’s alpha = 0.798).

Basic descriptive statistical methods such as frequencies, percentages, means, standard deviations, and cross-tabulations were conducted to summarize the data and determine the differences between groups for selected demographic variables. Descriptive statistics by level of government were summarized using frequency distribution tables. Preventive health measures scores were compared according to demographic characteristics with independent samples *t-*test, or one-way analysis of variance (ANOVA) test using Tukey HSD post-hoc comparison as appropriate. Bivariate and multivariable binary logistic regression analyses were used to identify factors associated with outcome variables. Odds ratios (ORs) and their 95% confidence intervals (CIs) for each predictor were estimated from the logistic regression to quantify the associations between potential predictors and outcome variables. The statistical significance level was set at p*<*0.05.

### Ethical approval

The study was approved by the Institutional Review Board of the College of Health Sciences at Addis Ababa University (AAU) (protocol number: 042/20/SPH). Informed consent was obtained from each participant. Permission to undertake this study was obtained from every relevant authority at all levels. Official letters from AAU were written to each institution to cooperate and participate in the survey. Written or oral prior to responding to the questions. Participation was voluntary, anonymous and any participant could withdraw from completing the questionnaire at any time and were at liberty to not answer any question they did not want to answer. Anonymity and data confidentiality were ensured. All personnel involved in the survey received orientation on COVID-19 infection prevention and control measures, and wore protective face masks and used sanitizers during data collection.

## Results

### Sociodemographic characteristics

In total, 1,730 eligible participants from 46 government institutions were invited to participate in the study and 1,577 participants completed the questionnaires. Of these, 1,573 were valid and used for analysis (91.6%). The completed questionnaires per institution ranged from 18-58, with an average of 34.3. About 91% of the study participants provided written informed consent, while 9% provided verbal informed consent. Table 1 shows the sociodemographic characteristics of the study participants. About 40% of the study participants were drawn from national institutions, 38.8% from Addis Ababa city administration institutions and 21.6% from Oromia Regional State institutions located in Addis Ababa. The majority of the respondents were in the age group of 18 and 39 years (73.3%), male (64.2%), with a bachelor’s degree or above (88.3%) and lived in Addis Ababa (82.2%). The mean (±SD) year of service in the institution was 6.6 (±6.4) years. About 19% of the respondents reported that they were tested for COVID-19, 7.1% reported any chronic illness, and only 2% were quarantined due to COVID-19.

**Table 1.** Characteristics of study participants by level of government

| Characteristics | Government level, n (%) |  |  | Total, n (%) |
| --- | --- | --- | --- | --- |
|  | National | Addis Ababa | Oromia |  |
| Gender |  |  |  |  |
| Male | 405 (64.9) | 350 (57.4) | 228 (67.3) | 983 (62.5) |
| Female | 206 (33.0) | 244 (40) | 99 (29.2) | 549 (34.9) |
| Unknown* | 13 (2.1) | 16 (2.6) | 12 (3.5) | 41 (2.6) |
| Age group (years) |  |  |  |  |
| 18-29 | 174 (27.9) | 175 (28.7) | 48 (14.2) | 397 (25.2) |
| 30-39 | 253 (40.5) | 258 (42.3) | 145 (42.8) | 656 (41.7) |
| 40-49 | 90 (14.4) | 94 (15.4) | 77 (22.7) | 261 (16.6) |
| ≥50 | 55 (8.8) | 32 (5.2) | 35 (10.3) | 122 (7.8) |
| Unknown | 52 (8.3) | 51 (8.4) | 34 (10.0) | 137 (8.7) |
| Level of education |  |  |  |  |
| ≤12<sup>th</sup> grade | 17 (2.7) | 23 (3.8) | 7 (2.1) | 47 (3.0) |
| Diploma** | 53 (8.5) | 65 (10.7) | 14 (4.1) | 132 (8.4) |
| Bachelor's degree | 318 (51.0) | 391 (64.1) | 180 (53.1) | 889 (56.5) |
| Master's degree or above | 226 (36.2) | 126 (20.7) | 121 (35.7) | 473 (30.1) |
| Unknown | 10 (1.6) | 5 (0.8) | 17 (5.0) | 32 (2.0) |
| Year of experience in the institution |  |  |  |  |
| <5 | 352 (56.4) | 228 (53.8) | 82 (24.2) | 762 (48.4) |
| 5-9 | 160 (25.6) | 167 (27.4) | 79 (23.3) | 406 (25.8) |
| 10-14 | 44 (7.1) | 61 (10.0) | 76 (22.4) | 181 (11.5) |
| ≥15 | 51 (8.2) | 39 (6.4) | 78 (23.0) | 168 (10.7) |
| Unknown | 17 (2.7) | 15 (2.5) | 24 (7.1) | 56 (3.6) |
| Household size |  |  |  |  |
| 1-3 | 267 (42.8) | 227 (37.2) | 89 (26.3) | 583 (37.1) |
| 4-5 | 231 (37.0) | 249 (40.8) | 142 (41.9) | 622 (39.5) |
| 6-7 | 76 (12.2) | 85 (13.9) | 65 (19.2) | 226 (14.4) |
| ≥8 | 24 (3.8) | 34 (5.6) | 17 (5.0) | 75 (4.8) |
| Unknown | 26 (4.2) | 15 (2.5) | 26 (7.7) | 67 (4.3) |
| Area of residence |  |  |  |  |
| Addis Ababa city | 580 (92.9) | 584 (95.7) | 129 (38.1) | 1293 (82.2) |
| Out of Addis Ababa | 37 (5.9) | 25 (4.1) | 147 (43.3) | 209 (13.3) |
| Unknown | 7 (1.1) | 1 (0.2) | 63 (18.6) | 71 (4.5) |
| Tested for COVID-19 |  |  |  |  |
| Yes | 156 (25.0) | 118 (19.3) | 26 (7.7) | 300 (19.1) |
| No | 448 (71.8) | 476 (78.0) | 251 (74.0) | 1175 (74.7) |
| Unknown | 20 (3.2) | 16 (2.6) | 62 (18.3) | 98 (6.2) |
| Reported any chronic illness |  |  |  |  |
| Yes | 35 (5.6) | 44 (7.2) | 33 (9.7) | 112 (7.1) |
| No | 483 (77.4) | 441 (72.3) | 232 (68.4) | 1156 (73.5) |
| Don't know or unknown | 106 (17.0) | 125 (20.5) | 74 (21.8) | 305 (19.4) |
| Quarantined due to COVID-19 |  |  |  |  |
| Yes | 15 (2.4) | 8 (1.3) | 8 (2.4) | 31 (2.0) |
| No | 605 (97.0) | 592 (97.0) | 318 (93.8) | 1515 (96.3) |
| Unknown | 4 (0.6) | 10 (1.6) | 13 (3.8) | 27 (1.7) |
| **Total, n (%)** | **624 (100)** | **610 (100)** | **339 (100)** | **1,573 (100)** |

\*Non-response; \*\*12<sup>th</sup> grade complete and one or more years of training

### Social distancing and preventive practices

Respondents were asked to indicate the types of protective measures they applied to prevent contracting COVID-19. Figure 1 presents the proportions of respondents who reported positively to the 14 social distancing measures and preventive practices taken in response to COVID-19. Overall, more than 9 in 10 respondents (95.9%) reported wearing facemask, 95.6% reported that they consistently followed the recommendations from the authorities to combat COVID-19, 92% reported that they recommended the wearing of facemask for healthy people out of the healthcare setting, 94.5% avoided close contact with people including handshaking, 94.1% reported frequently washing hands with water and soap, 90.8% covered mouth/nose while coughing or sneezing, 90.7% avoided touching eyes, nose and mouth (90.7%), and 89.5% practiced physical distancing. The majority of the respondents also reported avoiding mass gatherings and crowded places (88.1%), disinfected surfaces (77.6%), disinfected mobile phones (76.9%), restricted movement and traveling (71.8%), ate garlic, ginger and lemon (57.9%). The lowest level of compliance in response to COVID-19 was related to staying home, which was reported by 38.6% of participants.

**Figure 1.**
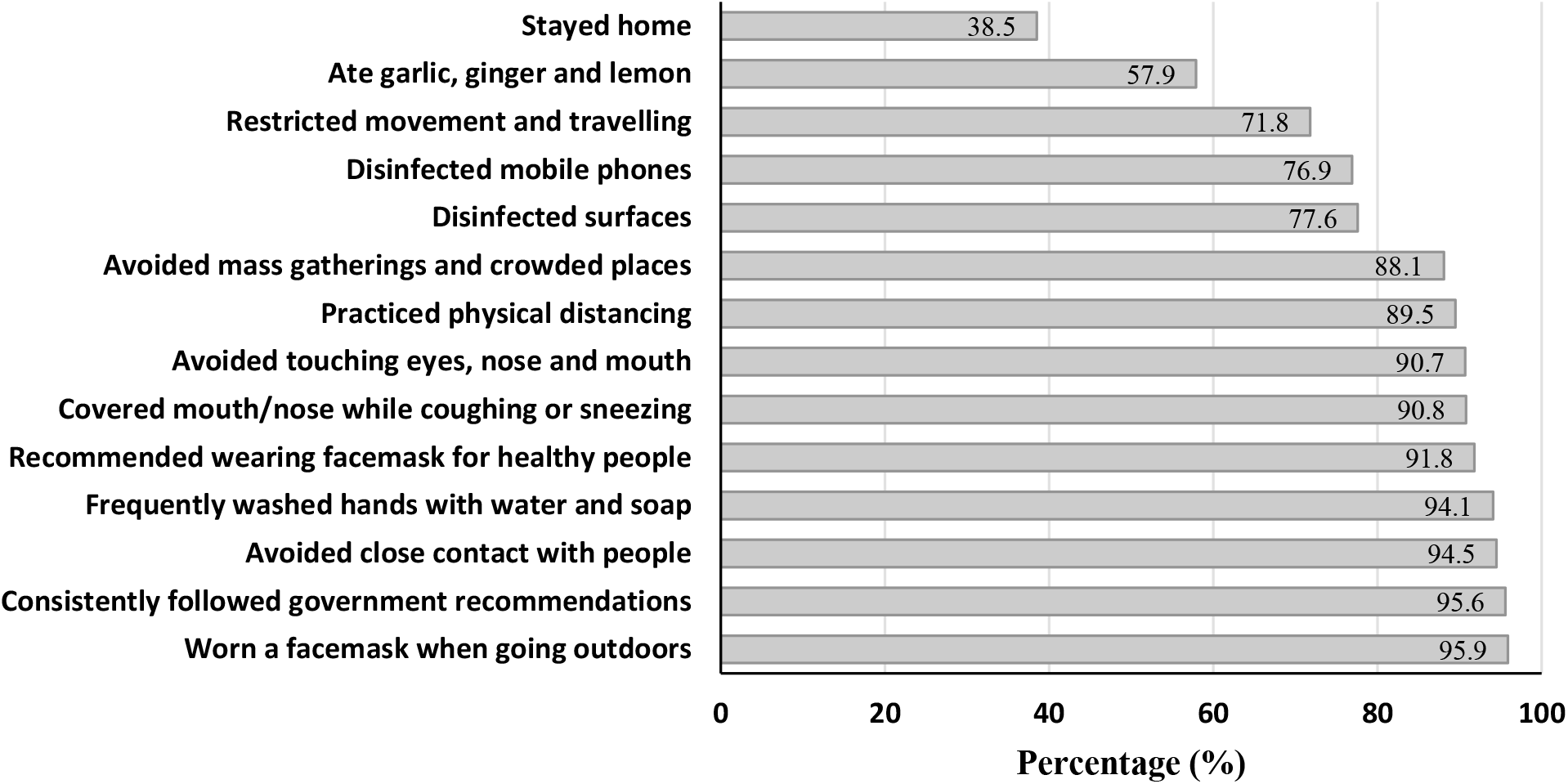
The proportions of respondents who reported positively to the 14 protective measures in response to COVID-19

Table 2 shows the distribution of the positive responses of the respondents to the 14 social distancing and preventive measures by the Government level. The majority (>90%) of the respondents reported the practice of wearing facemask, followed Government recommendations, avoided close contact with people, frequently washed hands and avoided touching eyes, nose and mouth across all the three Government levels. However, disinfecting surfaces (91.2%), staying home (45.7%), and restricting movement and travelling (84.1%) were more frequently reported in Oromia compared with respondents from Addis Ababa and national level. Whilst, wearing facemask (97.3%) and consistently following Government recommendations (97.1%) were more commonly reported among the national respondents than those from Oromia. This study also revealed that 64% of respondents from Oromia, 58% from national and 54% from Addis Ababa reported that they used garlic, ginger and lemon for prevention of coronavirus infection.

**Table 2.** Distribution of respondents reported preventive measures in response to COVID-19 by government level (n=14 items)

| Reported protective practices | Government level, % |  |  |
| --- | --- | --- | --- |
|  | National | Addis Ababa | Oromia |
| Worn a facemask when going outdoors | 97.3 | 95.6 | 94.1 |
| Consistently followed government recommendations | 97.1 | 95.1 | 93.8 |
| Avoided close contact with people and hand shaking | 95.7 | 94.8 | 92.0 |
| Frequently washed hands with water and soap | 95.4 | 94.1 | 91.7 |
| Recommended wearing facemask for healthy people | 93.9 | 93.0 | 85.8 |
| Covered mouth/nose while coughing or sneezing | 93.6 | 88.8 | 90.3 |
| Avoided touching eyes, nose and mouth | 92.0 | 89.7 | 90.0 |
| Practiced physical distancing | 90.9 | 88.0 | 89.7 |
| Avoided mass gatherings and crowded places | 91.8 | 86.9 | 83.5 |
| Disinfected surfaces | 75.5 | 72.1 | 91.2 |
| Disinfected mobile phones | 81.1 | 71.1 | 79.6 |
| Restricted movement and travelling | 71.3 | 65.4 | 84.1 |
| Ate garlic, ginger and lemon | 58.3 | 53.9 | 64.0 |
| Stayed home | 37.3 | 35.6 | 45.7 |
| *Total, n* | **624** | **610** | **339** |

The positive responses of the 14 social distancing and preventive practices in response to COVID-19 were added to produce the scores of the overall reported practice. Summing the positive responses across the total measures for each individual revealed a mean sample score of 11±2.3 measures taken and a median of 12 measures with an interquartile range (IQR) of 2, on a scale of 14. About 24% and 20% of the respondents scored 13 and 14 positive responses on the protective measures taken in response to COVID-19, respectively, while 10.6% scored 8 or less responses (Figure 2). Table 3 shows the distribution and comparison of the respondent’s mean scores of protective measures taken in response to COVID-19 by socio-demographic characteristics using ANOVA test and independent samples t-test. Female respondents had a statistically significant higher mean score responses (11.8±2.48) compared with males (p=0.007). Similarly, respondents who reported a household size of 4-5 had a significantly higher mean score responses (11.8±2.19) of protective practices compared with others (p=0.014). However, study participants from the Addis Ababa had a statistically significant lower mean score responses (11.2±2.59) compared with those from national and Oromia (p=0.001). There was no statistically significant difference in the mean score protective practices among the age groups, level of education, year of experience, area of residence, reported COVID-19 testing, whether quarantined or reporting any chronic illness.

**Table 3.**
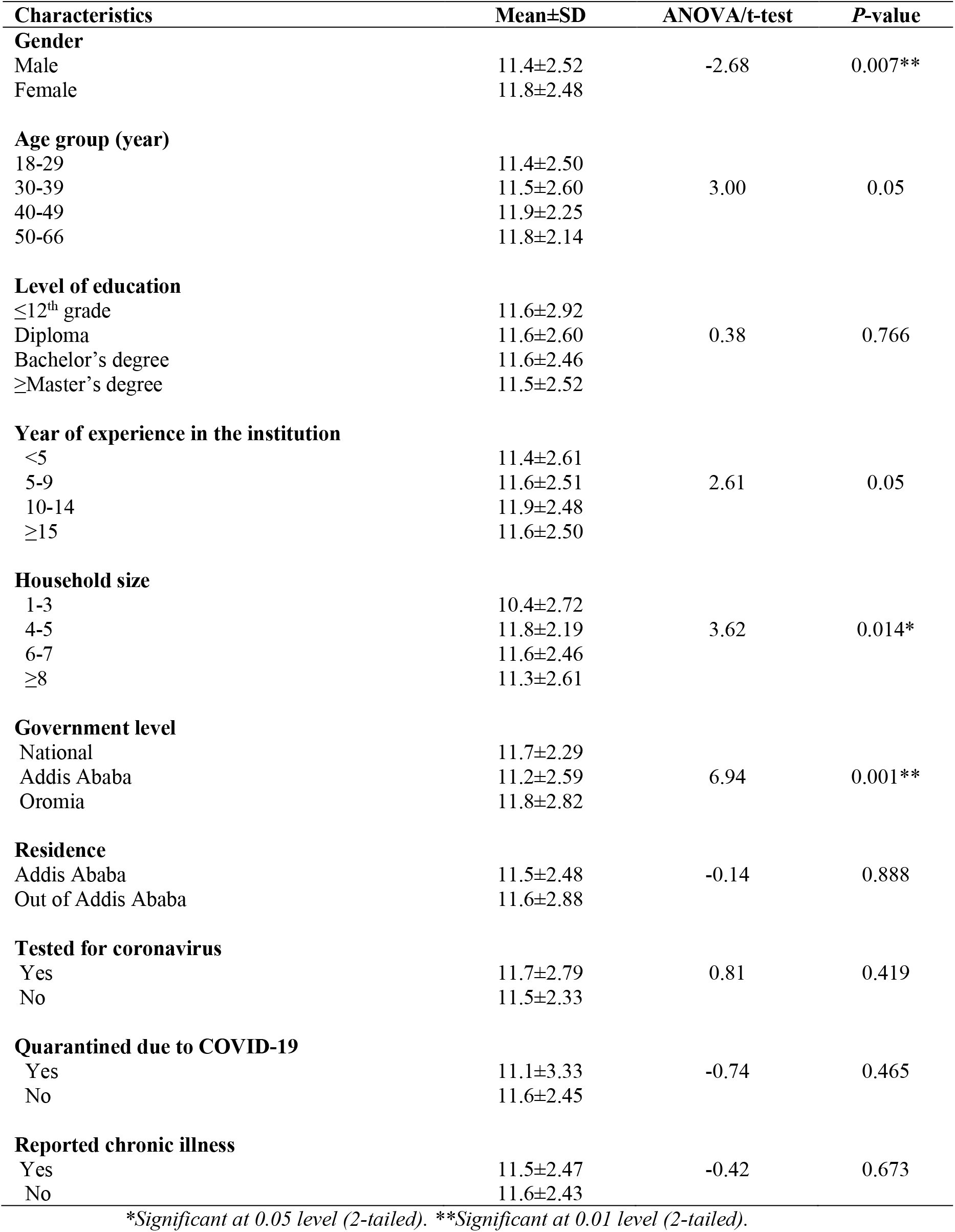
Comparison of protective measures mean score responses to COVID-19 between groups (n=14 items)

| Characteristics | Mean±SD | ANOVA/t-test | P-value |
| --- | --- | --- | --- |
| **Gender** |  |  |  |
| Male | 11.4±2.52 | -2.68 | 0.007** |
| Female | 11.8±2.48 |  |  |
| **Age group (year)** |  |  |  |
| 18-29 | 11.4±2.50 | 3.00 | 0.05 |
| 30-39 | 11.5±2.60 |  |  |
| 40-49 | 11.9±2.25 |  |  |
| 50-66 | 11.8±2.14 |  |  |
| **Level of education** |  |  |  |
| ≤12<sup>th</sup> grade | 11.6±2.92 | 0.38 | 0.766 |
| Diploma | 11.6±2.60 |  |  |
| Bachelor's degree | 11.6±2.46 |  |  |
| ≥Master's degree | 11.5±2.52 |  |  |
| **Year of experience in the institution** |  |  |  |
| <5 | 11.4±2.61 | 2.61 | 0.05 |
| 5-9 | 11.6±2.51 |  |  |
| 10-14 | 11.9±2.48 |  |  |
| ≥15 | 11.6±2.50 |  |  |
| **Household size** |  |  |  |
| 1-3 | 10.4±2.72 | 3.62 | 0.014* |
| 4-5 | 11.8±2.19 |  |  |
| 6-7 | 11.6±2.46 |  |  |
| ≥8 | 11.3±2.61 |  |  |
| **Government level** |  |  |  |
| National | 11.7±2.29 | 6.94 | 0.001** |
| Addis Ababa | 11.2±2.59 |  |  |
| Oromia | 11.8±2.82 |  |  |
| **Residence** |  |  |  |
| Addis Ababa | 11.5±2.48 | -0.14 | 0.888 |
| Out of Addis Ababa | 11.6±2.88 |  |  |
| **Tested for coronavirus** |  |  |  |
| Yes | 11.7±2.79 | 0.81 | 0.419 |
| No | 11.5±2.33 |  |  |
| **Quarantined due to COVID-19** |  |  |  |
| Yes | 11.1±3.33 | -0.74 | 0.465 |
| No | 11.6±2.45 |  |  |
| **Reported chronic illness** |  |  |  |
| Yes | 11.5±2.47 | -0.42 | 0.673 |
| No | 11.6±2.43 |  |  |

\*Significant at 0.05 level (2-tailed). \*\*Significant at 0.01 level (2-tailed).

**Figure 2.**
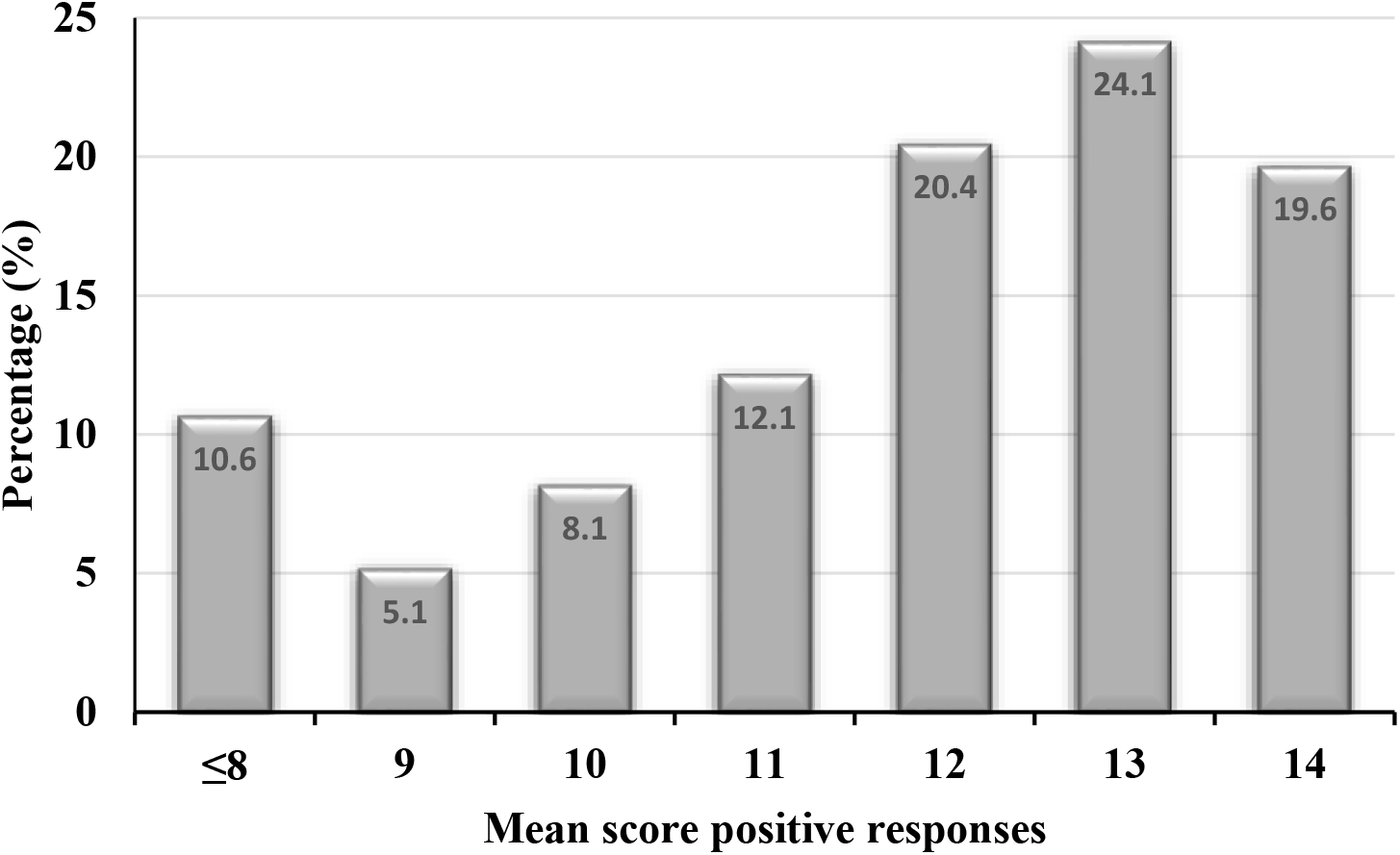
Mean score positive protective responses of the respondents in response to COVID-19

### Perceived effectiveness of mask wearing and associated factors

More than 80% of the participants perceived that consistently wearing a facemask is highly effective in preventing the transmission of coronavirus. Only 9.3% of the respondents disagreed or strongly disagreed about the effectiveness of facemask in the prevention of coronavirus infection. Nearly 21% of the respondents in Oromia neither agreed nor disagreed about the effectiveness of facemasks in the prevention of coronavirus infection. Table 4 shows the findings of the bivariate and multivariable logistic regression analyses of predictors associated with the respondent’s perceived effectiveness of consistently wearing facemask in preventing the infection due to coronavirus. The variables were initially assessed by bivariate analysis and retained in the multivariate models whether or not they were statistically significant. Among ten predictors assessed by bivariate logistic regression analyses, respondents who served in the institution more than 15 years (OR=0.68, 95% CI:0.45-1.03) and who were from Oromia (OR=0.31, 95% CI:0.22-0.43) perceived less effectiveness of facemask, whilst, study participants who resided in Addis Ababa (OR=2.02, 95% CI:1.44-2.83) and who were tested for coronavirus (OR=1.43, 95% CI:1.00-2.04) were more likely perceived higher effectiveness of facemasks to prevent coronavirus infection. In the multivariable logistic regression, participants from Oromia, compared to the national respondents, reported statistically significantly lower odds (adjusted OR=0.27, 95% CI:0.17-0.45) of perceived effectiveness of facemask in preventing coronavirus infection. However, the other predictor variables were not statistically significant in the multivariate analyses.

**Table 4.** Factors associated with perceived effectiveness of facemask in preventing coronavirus infection using logistic regression analyses

| Predictor | Total | Highly effective (%) | Crude<br>OR (95% CI)* | P-value | Adjusted<br>OR (95% CI) | P-value |
| --- | --- | --- | --- | --- | --- | --- |
| **Gender** |  |  |  |  |  |  |
| Male | 969 | 80.7 | 0.85 (0.65-1.12) | 0.251 | 0.75 (0.52-1.08) | 0.117 |
| Female | 544 | 83.1 | 1** |  | 1 |  |
| **Age group (year)** |  |  |  |  |  |  |
| 18-29 | 395 | 81.3 | 1 |  | 1 |  |
| 30-39 | 649 | 82.9 | 1.12 (0.81-1.55) | 0.503 | 1.24 (0.82-1.88) | 0.307 |
| 40-49 | 256 | 78.9 | 0.86 (0.58-1.28) | 0.460 | 0.96 (0.56-1.63) | 0.868 |
| 50-66 | 119 | 83.2 | 1.14 (0.66-1.96) | 0.634 | 1.40 (0.68-2.88) | 0.365 |
| **Education** |  |  |  |  |  |  |
| ≤12<sup>th</sup> grade | 47 | 89.4 | 1 |  | 1 |  |
| Diploma | 131 | 79.4 | 0.46 (0.17-1.27) | 0.134 | 0.41 (0.11-1.58) | 0.196 |
| Bachelor's degree | 879 | 80.0 | 0.48 (0.19-1.22) | 0.122 | 0.63 (0.18-2.19) | 0.463 |
| ≥Master's degree | 465 | 84.3 | 0.64 (0.25-1.66) | 0.639 | 0.91 (0.25-3.28) | 0.885 |
| **Experience (year)** |  |  |  |  |  |  |
| <5 | 754 | 83.3 | 1 |  | 1 |  |
| 5-9 | 401 | 81.0 | 0.86 (0.63-1.18) | 0.340 | 0.83 (0.57-1.22) | 0.351 |
| 10-14 | 180 | 81.7 | 0.89 (0.59-1.37) | 0.603 | 1.25 (0.72-2.18) | 0.436 |
| ≥15 | 162 | 77.2 | 0.68 (0.45-1.03) | 0.065 | 0.85(0.46-1.57) | 0.613 |
| **Household size** |  |  |  |  |  |  |
| 1-3 | 577 | 81.6 | 1 |  | 1 |  |
| 4-5 | 616 | 82.8 | 1.08 (0.81-1.46) | 0.600 | 1.30 (0.91-1.87) | 0.149 |
| 6-7 | 219 | 80.8 | 0.95 (0.64-1.41) | 0.794 | 1.06 (0.65-1.72) | 0.820 |
| ≥8 | 74 | 79.7 | 0.89 (0.48-1.62) | 0.693 | 1.19 (0.57-2.49) | 0.647 |
| **Government level** |  |  |  |  |  |  |
| National | 621 | 87.0 | 1 |  | 1 |  |
| Addis Ababa | 603 | 83.6 | 0.76 (0.56-1.05) | 0.096 | 0.80 (0.56-1.15) | 0.234 |
| Oromia | 328 | 67.4 | 0.31 (0.22-0.43) | <0.001 | 0.27 (0.17-0.45) | <0.001 |
| **Residence** |  |  |  |  |  |  |
| Addis Ababa | 1278 | 83.7 | 2.02 (1.44-2.83) | <0.001 | 1.04 (0.63-1.71) | 0.885 |
| Out of Addis Ababa | 206 | 71.8 | 1 |  | 1 |  |
| **Tested for COVID-19** |  |  |  |  |  |  |
| Yes | 297 | 85.9 | 1.43 (1.00-2.04) | 0.052 | 1.47 (0.95-2.27) | 0.082 |
| No | 1162 | 81.0 | 1 |  | 1 |  |
| **Reported any chronic illness** |  |  |  |  |  |  |
| Yes | 110 | 81.0 | 1.03 (0.62-1.70) | 0.925 | 1.18 (0.64-2.17) | 0.597 |
| No or didn't know | 1375 | 81.5 | 1 |  | 1 |  |
| **Quarantined due to COVID-19** |  |  |  |  |  |  |
| Yes | 31 | 77.4 | 0.77 (0.33-1.81) | 0.555 | 0.57 (0.17-1.92) | 0.365 |
| No | 1499 | 81.6 | 1 |  | 1 |  |

\*OR=Odds Ratio, CI=Confidence Interval; \*\*Reference

### COVID-19 testing and associated factors

About 19% (n=300) of the respondents reported that they had ever tested for coronavirus infection, with 25% of respondents at national level, 19.3% from Addis Ababa and 7.7% from Oromia. With regard to the question on the certainty of getting a COVID-19 test if needed, 11% of the respondents were completely sure, 18.1% were very sure, and 26% were somewhat sure. However, 15.1% did not know and 14.6% were not at all sure where and when to get coronavirus test if they wanted to be tested. Table 5 shows the results of the bivariate and multivariable logistic regression analyses conducted to explore factors associated with testing for COVID-19. In the bivariate analyses gender, age, year of experience, level of government, area of residence, reported chronic illness and being quarantined were significantly associated with testing for COVID-19. In the multivariable logistic regression analyses, the age groups 18-29 were more likely to test for coronavirus than the older age groups. In contrast, respondents from Oromia were less likely to test for coronavirus (adjusted OR=0.31, 95% CI:0.16-0.60) than those from national level. Gender, educational status, year of experience, household size, residence, reported chronic illness and being quarantined did not appear statistically significant in the multivariable logistic regression model to predict the odds of testing for COVID-19.

**Table 5.**
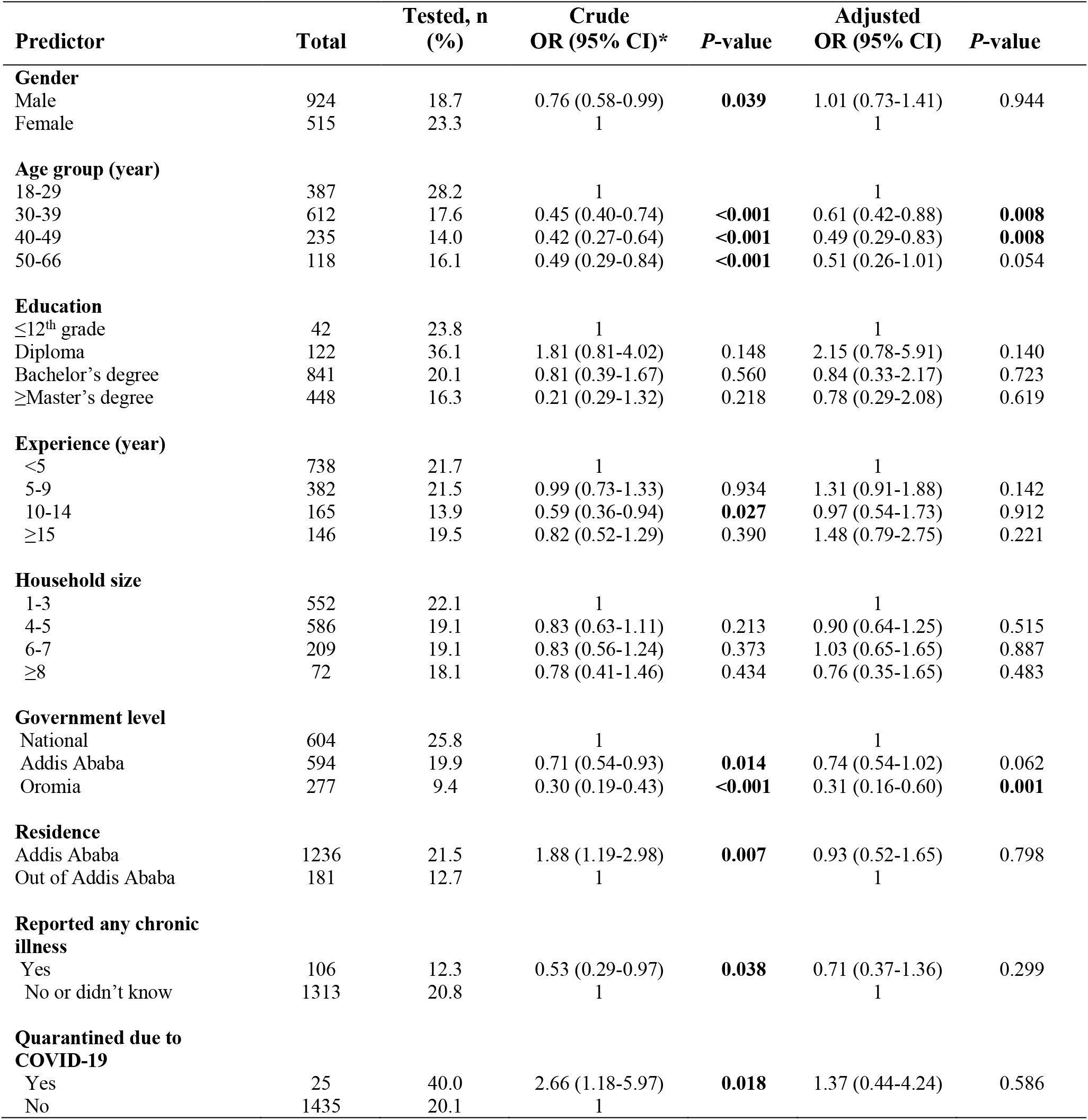
Factors associated with coronavirus testing in the study population using multiple logistic regression analyses

| Predictor | Total | Tested, n (%) | Crude OR (95% CI)* | P-value | Adjusted OR (95% CI) | P-value |
| --- | --- | --- | --- | --- | --- | --- |
| **Gender** |  |  |  |  |  |  |
| Male | 924 | 18.7 | 0.76 (0.58-0.99) | **0.039** | 1.01 (0.73-1.41) | 0.944 |
| Female | 515 | 23.3 | 1 |  | 1 |  |
| **Age group (year)** |  |  |  |  |  |  |
| 18-29 | 387 | 28.2 | 1 |  | 1 |  |
| 30-39 | 612 | 17.6 | 0.45 (0.40-0.74) | **<0.001** | 0.61 (0.42-0.88) | **0.008** |
| 40-49 | 235 | 14.0 | 0.42 (0.27-0.64) | **<0.001** | 0.49 (0.29-0.83) | **0.008** |
| 50-66 | 118 | 16.1 | 0.49 (0.29-0.84) | **<0.001** | 0.51 (0.26-1.01) | 0.054 |
| **Education** |  |  |  |  |  |  |
| ≤12<sup>th</sup> grade | 42 | 23.8 | 1 |  | 1 |  |
| Diploma | 122 | 36.1 | 1.81 (0.81-4.02) | 0.148 | 2.15 (0.78-5.91) | 0.140 |
| Bachelor's degree | 841 | 20.1 | 0.81 (0.39-1.67) | 0.560 | 0.84 (0.33-2.17) | 0.723 |
| ≥Master's degree | 448 | 16.3 | 0.21 (0.29-1.32) | 0.218 | 0.78 (0.29-2.08) | 0.619 |
| **Experience (year)** |  |  |  |  |  |  |
| <5 | 738 | 21.7 | 1 |  | 1 |  |
| 5-9 | 382 | 21.5 | 0.99 (0.73-1.33) | 0.934 | 1.31 (0.91-1.88) | 0.142 |
| 10-14 | 165 | 13.9 | 0.59 (0.36-0.94) | **0.027** | 0.97 (0.54-1.73) | 0.912 |
| ≥15 | 146 | 19.5 | 0.82 (0.52-1.29) | 0.390 | 1.48 (0.79-2.75) | 0.221 |
| **Household size** |  |  |  |  |  |  |
| 1-3 | 552 | 22.1 | 1 |  | 1 |  |
| 4-5 | 586 | 19.1 | 0.83 (0.63-1.11) | 0.213 | 0.90 (0.64-1.25) | 0.515 |
| 6-7 | 209 | 19.1 | 0.83 (0.56-1.24) | 0.373 | 1.03 (0.65-1.65) | 0.887 |
| ≥8 | 72 | 18.1 | 0.78 (0.41-1.46) | 0.434 | 0.76 (0.35-1.65) | 0.483 |
| **Government level** |  |  |  |  |  |  |
| National | 604 | 25.8 | 1 |  | 1 |  |
| Addis Ababa | 594 | 19.9 | 0.71 (0.54-0.93) | **0.014** | 0.74 (0.54-1.02) | 0.062 |
| Oromia | 277 | 9.4 | 0.30 (0.19-0.43) | **<0.001** | 0.31 (0.16-0.60) | **0.001** |
| **Residence** |  |  |  |  |  |  |
| Addis Ababa | 1236 | 21.5 | 1.88 (1.19-2.98) | **0.007** | 0.93 (0.52-1.65) | 0.798 |
| Out of Addis Ababa | 181 | 12.7 | 1 |  | 1 |  |
| **Reported any chronic illness** |  |  |  |  |  |  |
| Yes | 106 | 12.3 | 0.53 (0.29-0.97) | **0.038** | 0.71 (0.37-1.36) | 0.299 |
| No or didn't know | 1313 | 20.8 | 1 |  | 1 |  |
| **Quarantined due to COVID-19** |  |  |  |  |  |  |
| Yes | 25 | 40.0 | 2.66 (1.18-5.97) | **0.018** | 1.37 (0.44-4.24) | 0.586 |
| No | 1435 | 20.1 | 1 |  |  |  |

### Perceived adequacy of policy responses

Table 6 shows the perceptions of the respondents towards the policy decisions made by the Government to contain the spread of COVID-19 pandemic. Just under the third (31.3%) of the respondents strongly agreed that the policy responses that the Government had taken to contain the spread of coronavirus were fair and reasonable, and 38.5% agreed with the policy responses. However, 22.8% of the respondents in Oromia disagreed about the fairness and reasonability of policy responses taken by the Government. Over half (57.1%) of the study participants perceived that the current policy measures taken by the Government to contain the spread of coronavirus are inadequate (37.7%) or very inadequate (19.4%). More respondents from Oromia (63.7%) as compared with 59.3% in Addis Ababa and just about half (51.1%) at national level perceived that the current policy measures taken by the Government to contain the spread of coronavirus transmission were inadequate.

**Table 6.** Perceptions about policy responses to contain the spread of coronavirus (n=1573)

| Variable | Government level, % |  |  | Total, % |
| --- | --- | --- | --- | --- |
|  | National | Addis Ababa | Oromia |  |
| Policy responses that have been made by the Government to contain the spread of coronavirus are fair and reasonable |  |  |  |  |
| Strongly agree | 32.1 | 29.8 | 32.7 | 31.3 |
| Agree | 42.1 | 41.3 | 26.8 | 38.5 |
| Neither agree nor disagree | 11.5 | 9.2 | 13.9 | 11.1 |
| Disagree | 6.6 | 10.8 | 8.6 | 8.6 |
| Strongly disagree | 6.6 | 7.7 | 14.2 | 8.6 |
| Unknown* | 1.1 | 1.1 | 3.8 | 1.7 |
| What do you think about the adequacy of current measures by the Government to contain the spread of coronavirus? |  |  |  |  |
| Very adequate | 4.5 | 5.4 | 4.7 | 4.9 |
| Adequate | 21.5 | 12.0 | 17.1 | 16.8 |
| Neither adequate nor inadequate | 21.6 | 20.8 | 10.6 | 18.9 |
| Inadequate | 34.3 | 38.2 | 43.1 | 37.7 |
| Very inadequate | 16.8 | 21.3 | 20.6 | 19.4 |
| Unknown* | 1.3 | 2.3 | 3.8 | 2.2 |
| Total, n | 624 | 610 | 339 | 1573 |

\*Non-response

## Discussion

The findings of the current study revealed high level of reported practices of COVID-19 protective measures, particularly with regard to mask wearing in public (95.9%), avoiding close contact with people and handshaking (94.5%), frequent handwashing with water and soap (94.1%), maintaining physical distancing (89.5%), avoiding crowds and gatherings (88%), and movement restriction (72%). A study conducted during the early phase of the pandemic in Ethiopia reported a lower level of protective behaviors against the COVID-19 infection such as washing hands frequently (77%), avoiding shaking hands (54%) and not going to crowded places (33%) [30]. A study conducted in Malaysia reported that a high proportion of respondents already adopted avoidance behaviors such as not going to crowded places (83%) and practicing proper hand hygiene (88%) at the time of their study in late March 2020, despite low level of mask wearing (51%) [35]. It has been observed that staying at home, maintaining physical distancing, avoiding public transport and escaping going to public places is particularly difficult for many government employees, resulting in less adoption of these protective measures.

The current study analyzed factors affecting the mean score of the practice of social distancing and preventive practices of COVID-19. The study indicates that female gender was more likely than men to report social distancing measures and preventive practices probably since they had greater motivation for health or perceived susceptibility to COVID-19 than men. The results are consistent with those reported during the pandemic [32]. In a cross-sectional study of preventive health behaviors in Iran, the mean score of preventive behaviors from COVID-19 was higher in females than males [36]. In studies carried out on the pandemic of SARS (H1N1) epidemic in Hong Kong and Singapore, women were more likely than men to adopt protective behaviors such as handwashing, mask wearing, and respiratory hygiene [37-39]. Moreover, respondents working in Oromia or national offices and those having 4-5 household size were more likely than others to report protective health measures against COVID-19 probably due to the difference in their awareness and knowledge levels of the disease.

A study conducted during the early stage of the COVID-19 epidemic in China identified that respondents adopted important protective behaviors, and nearly all study participants (98%) wore masks when going out in public during the study period [40]. In Hong Kong, individual behaviors in the population changed in response to the threat of COVID-19, and 85% of respondents reported avoiding crowded places and 99% reported wearing facemasks in public [16]. Until effective treatments and vaccines are available, behavioral interventions such as social distancing and preventive practices are the most recommended tools to prevent and control the spread of COVID-19 [41]. Studies found that the non-pharmaceutical interventions (including border restrictions, quarantine, isolation, physical distancing, and changes in population behavior) were substantially associated with reduced transmission of COVID-19 [16]. Maintaining and sustaining high levels of actual protective practices, particularly wearing mask at public, is critically important and concerted efforts should be made by the government, media, healthcare professionals, local organizations, the community and individuals to combat COVID-19 focusing on preventive health behaviors.

According to the current study, about 80% of the respondents perceived that consistently wearing facemask is highly effective in preventing the spread of coronavirus, and 92% supported its use by healthy people in public. At the time of data collection for this study, discussions were underway whether facemasks should be used in public by healthy people out of the healthcare setting. The WHO earlier in April advised not to use facemasks in the community setting by healthy individuals without respiratory symptoms [14], but later recommended universal masking in June [15] when asymptomatic and pre-symptomatic infectiousness of SARS-CoV-2 was established [42, 43]. Recently, masks have mainly been worn by individuals in the general community who have certain respiratory symptoms and by those who feel particularly susceptible to infection and want to protect themselves [16].

Studies from China and the US [19] have shown wearing masks is effective in reducing the risk of infection and mitigating the spread of COVID-19, particularly when combined with other preventive measures such as physical distancing and frequent handwashing [19, 44]. A recent systematic review and meta-analysis study funded by the WHO demonstrated the effectiveness of physical distancing of 1m or more and the use of facemasks in public and health-care settings in the prevention of coronavirus transmission, where both interventions reduced the risk of infection of coronavirus by more than 80% [45]. A case-control study from Thailand found that mask wearing, frequent handwashing and social distancing of ≥1m were independently and significantly associated with reduced risk of SARS-CoV-2 infection among the general public [46]. Another study showed that the use of facemasks among the general population significantly reduced total infections and the number of deaths, and mask wearing is considered as one of the most effective public health measures in mitigating transmission of SARS-CoV-2 [47]. These studies provide the most reliable evidence on the effectiveness of simultaneous use of social distancing measures and wearing of facemasks at community level to mitigate the spread of COVID-19. However, there are still inconsistent scientific evidence that contradicts the effectiveness of using facemasks by healthy people in the community to prevent infection with SARS-CoV-2 [48].

The current study also assessed factors affecting the perceived effectiveness of mask wearing against the prevention of coronavirus infection. It was found that respondents from Oromia were less likely to report the effectiveness of wearing facemasks to prevent the transmission of SARS-CoV-2 than those from national or Addis Ababa levels, which could be associated with inadequate awareness or knowledge about the protective benefits of wearing facemask against the infection of SARS-COV-2. The more people became aware of the risk of COVID-19 to themselves, the more likely they begin practicing protective behaviors like mask wearing, handwashing and social distancing. In Ethiopia, mask wearing in public is not a common practice before the occurrence of COVID-19 pandemic. Even after the onset of the pandemic, mask wearing practice in public was very minimal, but it increased immediately when the mandatory policy of facemask wearing for all people in public and working places was enforced at the end of May 2020.

The present study also showed that respondents aged between 18 and 29 years were more likely to be tested for COVID-19 compared to the older respondents, while study participants from Oromia were less likely to be tested compared with respondents from national or Addis Ababa levels. Behavioral changes are currently one of the main tools to fight against COVID-19. These changes include practicing physical distancing, frequent handwashing, using hand sanitizers, wearing facemasks and testing for COVID-19. Nonetheless, these behavioral measures are effective if they are widely accepted and applied by the public. To have these measures widely understood and implemented by the community, the government needs to embark upon heightened mass campaigns to educate the public about the significance of frequent handwashing, wearing facemasks, and social distancing in containing the transmission of SARS-COV-2.

A significant proportion (58%) of respondents in the current study reported that they used garlic, ginger and lemon to protect themselves against SARS-COV-2 infection, which indicates unconfirmed practices or misconceptions. Although these home remedies are important ingredients of our daily food and may have some medicinal properties, it is a great misconception to believe and use them against COVID-19 since they have not been tested against SARS-COV-2. Studies have found no evidence that the use of herbal remedies such as garlic and ginger is effective against infection from coronavirus or cure from COVID-19 [49, 50]. The WHO has also confirmed that there is no evidence that eating garlic or ginger has protected people from SARS-COV-2 infection [51]. Current evidence shows that using ginger or garlic or combining them with other ingredients, such as lemon, or drinking hot ginger tea will not prevent or cure COVID-19.

At the time of this study, the spread of COVID-19 in Addis Ababa city cumulatively increased from 1,625 on 8^th^ June to 2,988 on 19^th^ June, with an average of 114 per day. Despite efforts to contain and mitigate the transmission of coronavirus in the city, the virus has continued to spread to all parts of the city at an alarming rate and more cases from the community have continued to emerge on a daily basis. COVID-19 appeared to quickly spread in Ethiopia through the movement and frequent contact between people. Physical distancing has remained a major challenge due to overcrowding and, people are confronted with the logistical and communication problems particularly due to the shortage of means of transportation. Staying home approaches were particularly challenged in the context of poverty in the city where many residents lack adequate shelter, sanitation, and economic means for livelihood. Although staying home and physical distancing slows the transmission of SARS-COV-2, they result in heavy toll particularly on the informal economic and casual labor sector due to search of income for the day-to-day livelihood [24].

Ethiopia declared a state of emergency in April to mitigate the spread of COVID-19 pandemic [25]. Mandatory facemask wearing at banks, marketplaces, transport depots, in public transit, shops, pharmacies, places where public services are provided or any other public space of mass gatherings was mandated in the state of emergency. In addition, the Government also made mandatory facemask wearing for all people outside of their homes or offices on 27^th^ May. As a result, the practice of social distancing measures and preventive behaviors such as mask wearing in public have been significantly improved until the state of emergency was lifted on 11^th^ September 2020. Unfortunately, this was followed by the roll back of the already adopted social distancing measures and preventive practices by the public. Consequently, the EPHI adopted a directive on 5^th^ October, which enforced mandatory wearing of facemasks in public places, maintaining physical distancing of at least 2 m apart from other people; regular handwashing with soap or alcoholic-based sanitizers; and prohibited any organization to provide service to any person who is not wearing a facemask [52]. However, these measures have not been well enforced and the public has become reluctant regarding the social distancing and preventive practices of COVID-19. As the practice of social distancing involves staying home and away from others as much as possible to help prevent spread of COVID-19, Ethiopia mainly promoted physical distancing which involves the need to stay at least 2 m from others, complemented by wearing facemasks. Studies have shown the effectiveness of social distancing and mandatory facemask in public in mitigating the spread of COVID-19 in many countries, and both interventions and the simultaneous implementation of other preventive measures have been identified as the strategic priorities for containing COVID-19 [53].

### Limitations

This study had some limitations including selection bias that deserve explanations. First, the study only included government employees in Addis Ababa, and it failed to include unemployed people or other individuals working in non-governmental or private institutions, leading to concerns about the representativeness of the sample. There might be differences in adapting protective health measures between employed and unemployed people as well as between employees of governmental and non-governmental institutions. Second, due to the threat of COVID-19 and the physical distancing rule, it was not possible to conduct either a face-to-face interview or a community-based representative study. Third, the data presented in this study are based on retrospective self-reports of respondents without verification, thus the results might be subject to social desirability and recall biases.

### Conclusions

Despite the limitations, this study generated valuable information about protective behaviors of COVID-19 among government employees. The findings showed higher social distancing and preventive practices in response to COVID-19. In the current pandemic scenario, people should follow the Governments’ instructions and properly apply social distancing measures, wearing facemasks, and washing hands frequently with water and soap. Rules and regulations imposed by the Government should be properly enforced in order to control the pandemic. The findings have significant implications in identifying ways of promoting compliance with recommended protective health behaviors to effectively control the ongoing COVID-19 pandemic. The results of this study can be used as a baseline data to the government and researchers for other larger studies to identify factors significantly associated with preventive health measures.

## Data Availability

Data will be available based on reasonable request to the corresponding author

## Abbreviations

ANOVA: Analysis of variance
AAU: Addis Ababa University
COVID-19: Coronavirus disease 2019
CSPro: Census Surveys Professional
EPHI: Ethiopian Public Health Institute
HSD: Honestly significant difference
IRB: Institutional Review Board
MoH: Ministry of Health
SARS-CoV-2: Severe acute respiratory syndrome coronavirus 2
SD: Standard deviation
SPH: School of Public Health
SPSS: Statistical Package for Social Sciences
WHO: World Health Organization

## Ethical approval

The study was approved by the Institutional Review Board of the College of Health Sciences at Addis Ababa University (AAU) (protocol number: 042/20/SPH). Informed consent was obtained from each participant.

## Author Contributions

WD, AW, WAA and WA conceptualized and designed the study. SG, WD and AW supervised the field data collection. WD prepared the final dataset for analysis. WD and AW analyzed and interpreted the data. WD drafted the manuscript. AW provided intellectual role in improving the manuscript. AW, SG, WAA and WA provided major roles in revising the manuscript. All authors read and approved the final manuscript.

## Consent for publication

Not applicable

## Availability of data and materials

The datasets used and analyzed during the current study will be open access and available via an online repository.

## Acknowledgements

The authors are grateful to the research staff at the College of Health Sciences. The authors would also like to thank the data collectors and study participants for their time and contributing to the research.

---

**Additional file 1**: List of institutions/organizations included in the survey with collected samples, June 2020

**Additional file 2:** Questionnaires for assessment of social distancing and preventive practices of COVID-19 among government employees in Addis Ababa, June 2020

## References

1. Adhikari SP, Meng S, Wu YJ, Mao YP, Ye RX, Wang QZ, et al. Epidemiology, causes, clinical manifestation and diagnosis, prevention and control of coronavirus disease (COVID-19) during the early outbreak period: a scoping review. Infect Dis Poverty 2020; 9(1):29. Doi: 10.1186/s40249-020-00646-x.

2. Li Q, Guan X, Wu P, Wang X, Zhou L, Tong Y, et al. Early transmission dynamics in Wuhan, China, of novel coronavirus-infected pneumonia. N Engl J Med. 2020; 382(13):1199–1207. Doi: 10.1056/NEJMoa2001316.

3. Zhu N, Zhang D, Wang W, Li X, Yang B, Song J, et al. A Novel coronavirus from patients with pneumonia in China, 2019. N Engl J Med. 2020;382(8):727–733. Doi: 0.1056/NEJMoa2001017.

4. World Health Organization. Statement on the second meeting of the International Health Regulations (2005) Emergency Committee regarding the outbreak of novel coronavirus (2019-nCoV). 2020. https://www.who.int/news-room/detail/30-01-2020-statement-on-the-second-meeting-of-the-international-healthregulations-(2005)-emergency-committee-regarding-the-outbreak-of-novelcoronavirus-(2019-ncov). accessed 15 August 2020.

5. Alqahtani JS, Oyelade T, Aldhahir AM, Alghamdi SM, Almehmadi M, Alqahtani AS, et al. Prevalence, Severity and Mortality associated with COPD and Smoking in patients with COVID-19: A Rapid Systematic Review and Meta-Analysis. PLOS ONE. 2020; 15(5): e0233147. https://doi.org/10.1371/journal.pone.0233147 PMID: 32392262.

6. Albitar O, Ballouze R, Ooi JP, Sheikh Ghadzi SM. Risk factors for mortality among COVID-19 patients. Diabetes Res Clin Pract. 2020;166:108293. doi:10.1016/j.diabres.2020.108293.

7. Worldometers. COVID-19 Virus Pandemic (Live). https://www.worldometers.info/coronavirus/. 2020. Last updated: 14 December 2020, 05:33 GMT.

8. Gilbert M, Pullano G, Pinotti F, Valdano E, Poletto C, Boëlle PY, et al. Preparedness and vulnerability of African countries against importations of COVID-19: a modelling study. Lancet. 2020;395(10227):871–877. doi:10.1016/S0140-6736(20)30411-6.

9. Zelalem K, Sabit A, Firmaye B, Dagmawit S, Ermias W, Samson M, et al. Rapid Evidence synthesis on COVID-19 Pandemic to Inform Ethiopian Ministry of Health: Knowledge Translation Directorate, Ethiopian Public Health Institute, Addis Ababa, Ethiopia, April 2020.

10. Ethiopian Public Health Institute. National Public Health Emergency Operation Center. COVID-19 Pandemic Preparedness and Response. Bulletin No. 1, May 03, 2020. Addis Ababa, Ethiopia.

11. Ethiopian Public Health Institute. National Public Health Emergency Operation Center. COVID-19 Pandemic Preparedness and Response. Bulletin No. 6, June 08, 2020. Addis Ababa, Ethiopia.

12. Wilder-Smith A, Freedman DO. Isolation, quarantine, social distancing and community containment: pivotal role for old-style public health measures in the novel coronavirus (2019-nCoV) outbreak. J Travel Med. 2020; 27(2), taaa020, https://doi.org/10.1093/jtm/taaa020.

13. World Health Organization. Recommendations to Member States to improve hand hygiene practices to help prevent the transmission of the COVID-19 virus: interim guidance, 1 April 2020. World Health Organization, 2020. https://apps.who.int/iris/handle/10665/331661. accessed 15 September 2020.

14. World Health Organization. Considerations in adjusting public health and social measures in the context of COVID-19: interim guidance, 16 April 2020. https://www.who.int/publications/i/item/considerations-in-adjusting-public-health-and-social-measures-in-the-context-of-covid-19-interim-guidance.WorldHealthOrganization, 2020. Accessed 15 September 2020.

15. World Health Organization. Advice on the use of masks in the context of COVID-19: interim guidance, 5 June 2020. https://www.who.int/publications/i/item/advice-on-the-use-of-masks-in-the-community-during-home-care-and-in-healthcare-settings-in-thecontext-of-the-novel-coronavirus-(2019-ncov)-outbreak. World Health Organization, 2020. Accessed 15 September 2020.

16. Cowling BJ, Ali ST, Ng TWY, Tsang TK, Li JCM, Fong MW, et al. Impact assessment of non-pharmaceutical interventions against coronavirus disease 2019 and influenza in Hong Kong: an observational study. Lancet Public Health. 2020;5(5):e279–e288. Doi: 10.1016/S2468-2667(20)30090-6.

17. Gandhi M, Beyrer C, Goosby E. Masks Do More Than Protect Others During COVID-19: Reducing the Inoculum of SARS-CoV-2 to Protect the Wearer. J Gen Intern Med. 2020;35(10):3063–3066. Doi: 10.1007/s11606-020-06067-8.

18. McGrail DJ, Dai J, McAndrews KM, Kalluri R. Enacting national social distancing policies corresponds with dramatic reduction in COVID19 infection rates. PLOS ONE. 2020; 15(7):e0236619. https://doi.org/10.1371/journal.pone.0236619.

19. Wang Y, Tian H, Zhang L, Zhang M, Guo D, Wu W, et al., Reduction of secondary transmission of SARS-CoV-2 in households by face mask use, disinfection and social distancing: a cohort study in Beijing, China. BMJ Glob Health. 2020; 5(5):e002794. doi:10.1136/bmjgh-2020-002794.

20. Kim S, Ko Y, Kim Y-J, Jung E. The impact of social distancing and public behavior changes on COVID-19 transmission dynamics in the Republic of Korea. PLoS ONE. 2020; 15(9): e0238684. https://doi.org/10.1371/journal.pone.0238684.

21. Stutt Rojh, Retkute R, Bradley M, Gilligan CA, Colvin J. A modelling framework to assess the likely effectiveness of facemasks in combination with ‘lock-down’ in managing the COVID-19 pandemic. Proc R Soc. 2020; A 476: 20200376. http://dx.doi.org/10.1098/rspa.2020.0376.

22. Fung IC, Cairncross S. Effectiveness of handwashing in preventing SARS: a review. Trop Med Int Health. 2006;11(11):1749–58. Doi: 10.1111/j.1365-3156.2006.01734.x.

23. Suess T, Remschmidt C, Schink SB, Schweiger B, Nitsche A, Schroeder K, et al. The role of facemasks and hand hygiene in the prevention of influenza transmission in households: results from a cluster randomised trial; Berlin, Germany, 2009-2011. BMC Infect Dis. 2012;12:26. Doi: 10.1186/1471-2334-12-26.

24. Mehtar S, Preiser W, Lakhe NA, Bousso A, TamFum JM, Kallay O, et al. Limiting the spread of COVID-19 in Africa: one size mitigation strategies do not fit all countries. Lancet Glob Health. 2020;8(7):e881–e883. Doi: 10.1016/S2214-109X(20)30212-6.

25. The Federal Democratic Republic of Ethiopia. A Regulation Issued to Implement the State of Emergency Proclamation No. 3/2020 Enacted to Counter and Control the Spread of COVID-19 and Mitigate its Impact. Council of Ministers Regulation No. 466/2020. 20th April 2020. Addis Ababa, Ethiopia.

26. Ministry of Health. National Comprehensive COVID-19 Management Handbook. 1st Edition, April 2020. Addis Ababa, Ethiopia.

27. Clark C, Davila A, Regis M, Kraus S. Predictors of COVID-19 voluntary compliance behaviors: An international investigation. Glob Transit. 2020;2:76–82. Doi: 10.1016/j.glt.2020.06.003.

28. Aynalem YA, Akalu TY, Gebresellassie B, Sharew NT, Shiferaw WS. Assessment of undergraduate student knowledge, practices, and attitude towards COVID-19 in Debre Berhan University, Ethiopia. Research Square. 2020; DOI: 10.21203/rs.3.rs-28556/v1.

29. Bekele D, Tolossa T, Tsegaye R, Teshome W. The knowledge and practice towards COVID-19 pandemic prevention among residents of Ethiopia: An Online Cross-Sectional Study. 2020, bioRxiv preprint doi:https://doi.org/10.1101/2020.06.01.127381.

30. Kebede Y, Yitayih Y, Birhanu Z, Mekonen S, Ambelu A. Knowledge, perceptions and preventive practices towards COVID-19 early in the outbreak among Jimma university medical center visitors, Southwest Ethiopia. PLOS ONE 2020; 15(5): e0233744. https://doi.org/10.1371/journal.pone.0233744.

31. WHO. Survey tool and guidance: behavioral insights on COVID-19 WHO/EURO:2020-696-40431-54222. 29 July 2020. https://apps.who.int/iris/handle/10665/333549.

32. Bish A, Michie S. Demographic and attitudinal determinants of protective behaviours during a pandemic: a review. Br J Health Psychol. 2010; 15:797–824. DOI: 10.1348/135910710X485826.

33. Moran KR, Del Valle SY. A Meta-Analysis of the Association between Gender and Protective Behaviors in Response to Respiratory Epidemics and Pandemics. PLoS ONE 2016; 11(10): e0164541. DOI:10.1371/journal.pone.0164541.

34. Atchison CJ, Bowman L, Vrinten C, Redd R, Pristera D, Eaton JW, et al. Perceptions and behavioural responses of the general public during the COVID-19 pandemic: A cross-sectional survey of UK Adults. medRxiv; 2020. DOI: 10.1101/2020.04.01.20050039.

35. Azlan AA, Hamzah MR, Sern TJ, Ayub SH, Mohamad E. Public knowledge, attitudes and practices towards COVID-19: A cross-sectional study in Malaysia. PLOS ONE 2020; 15(5):e0233668. https://doi.org/10.1371/journal.pone.0233668.

36. Shahnazi H, Ahmadi-Livani M, Pahlavanzadeh B, Rajabi A, Hamrah MS, Charkazi A. Assessing preventive health behaviors from COVID-19: a cross sectional study with health belief model in Golestan Province, Northern of Iran. Infect Dis Poverty 2020; 9:157. https://doi.org/10.1186/s40249-020-00776-2.

37. Lau JTF, Yang X, Tsui H, Kim JH. Monitoring community responses to the SARS epidemic in Hong Kong: from day 10 to day 62. J Epidemiol Community Health 2003;57:864–870. doi:10.1136/jech.2003.017483.

38. Leung GM, Quah S, Ho LM, Ho SY, Hedley AJ, Lee HP, Lam TH. A tale of two cities: Community psychobehavioral surveillance in Hong Kong and Singapore during the severe acute respiratory syndrome epidemic. Infect Control Hosp Epidemiol. 2004; 25(12), 1033–1041.

39. Tang CS, Wong CY. Factors influencing the wearing of facemasks to prevent the severe acute respiratory syndrome among adult Chinese in Hong Kong. Prev Med. 2004; 39(6):1187–93. Doi: 10.1016/j.ypmed.2004.04.032.

40. Zhong BL, Luo W, Li HM, Zhang QQ, Liu XG, Li WT, Li Y. Knowledge, attitudes, and practices towards COVID-19 among Chinese residents during the rapid rise period of the COVID-19 outbreak: a quick online cross-sectional survey. Int J Biol Sci. 2020; 16(10): 1745–1752. doi:10.7150/ijbs.45221.

41. Allegrante JP, Auld ME, Natarajan S. Preventing COVID-19 and Its Sequela: “There Is No Magic Bullet… It’s Just Behaviors”. Am J Prev Med 2020;59(2):288–292.

42. Gandhi M, Yokoe DS, Havlir DV. Asymptomatic Transmission, the Achilles’ Heel of Current Strategies to Control Covid-19. N Engl J Med. 2020;382(22):2158–2160. doi:10.1056/NEJMe2009758.

43. Arons MM, Hatfield KM, Reddy SC, Kimball A, James A, Jacobs JR, et al. Presymptomatic SARS-CoV-2 Infections and Transmission in a Skilled Nursing Facility. N Engl J Med. 2020;382(22):2081–2090. doi:10.1056/NEJMoa2008457.

44. Lyu W, Wehby GL. Community Use of Face Masks And COVID-19: Evidence From A Natural Experiment of State Mandates in The US. Health Aff (Millwood). 2020;39(8):1419–1425. doi:10.1377/hlthaff.2020.00818.

45. Chu DK, Akl EA, Duda S, Solo K, Yaacoub S, Schünemann HJ, et al. COVID-19 Systematic Urgent Review Group Effort (SURGE) study authors. Physical distancing, face masks, and eye protection to prevent person-to-person transmission of SARS-CoV-2 and COVID-19: a systematic review and meta-analysis. Lancet. 2020;395(10242):1973–1987. doi:10.1016/S0140-6736(20)31142-9.

46. Doung-ngern P, Suphanchaimat R, Panjangampatthana A, Janekrongtham C, Ruampoom D, Daochaeng N, et al. Case-Control Study of Use of Personal Protective Measures and Risk for SARS-CoV 2 Infection, Thailand. Emerg Infect Dis. 2020;26(11):2607–2616. https://dx.doi.org/10.3201/eid2611.203003.

47. Worby CJ, Chang HH. Face mask use in the general population and optimal resource allocation during the COVID-19 pandemic. Nat Commun. 2020; 11:4049. https://doi.org/10.1038/s41467-020-17922-x.

48. Bundgaard H, Bundgaard JS, Raaschou-Pedersen Det, von Buchwald C, Todsen T, Norsk JB, et al. Effectiveness of Adding a Mask Recommendation to Other Public Health Measures to Prevent SARS-CoV-2 Infection in Danish Mask Wearers: A Randomized Controlled Trial. Ann Intern Med. 2020 Nov 18:M20–6817. doi:10.7326/M20-6817.

49. Barati F, Pouresmaieli M, Ekrami E, Asghari S, Ziarani FR, Mamoudifard M. Potential Drugs and Remedies for the Treatment of COVID-19: a Critical Review. Biol Proced Online 2020; 22:15. https://doi.org/10.1186/s12575-020-00129-1.

50. Phillips K. No, Bananas Don’t Cure HIV, Nor Will Garlic Cure COVID-19: Searching for, Assessing, and Consuming Health Information Online. J Consum Health Internet. 2020; 24:175–185, DOI: 10.1080/15398285.2020.1755149.

51. World Health Organization. Infographic. “Can eating garlic help prevent infection with the new coronavirus? [Online image]. WHO, EPI-WIN Mythbuster. 2020. https://www.who.int/emergencies/diseases/novel-coronavirus-2019/advice-for-public/myth-busters.

52. EPHI. A Directive Issued for the Prevention and Control of COVID-19 Pandemic. Directive No. 30/2020. October 05, 2020. Addis Ababa, Ethiopia.

53. Bo Y, Guo C, Lin C, Zeng Y, Li HB, Zhang Y, et al. Effectiveness of non-pharmaceutical interventions on COVID-19 transmission in 190 countries from 23 January to 13 April 2020. Int J Infect Dis. 2020;102:247–253. doi:10.1016/j.ijid.2020.10.066.

